# Influence of blood pressure on pneumonia risk: Epidemiological association and Mendelian randomisation in the UK Biobank

**DOI:** 10.1101/2020.04.19.20071936

**Authors:** Seyedeh M. Zekavat, Michael Honigberg, James Pirruccello, Puja Kohli, Elizabeth W. Karlson, Christopher Newton-Cheh, Hongyu Zhao, Pradeep Natarajan

## Abstract

**Objectives:** To determine whether elevated blood pressure influences risk for respiratory infection.

**Design:** Prospective, population-based epidemiological and Mendelian randomisation studies.

**Setting:** UK Biobank.

**Participants:** 377,143 self-identified British descent (54% women; median age 58 years) participants in the UK Biobank.

**Main outcome measures:** First incident pneumonia over an average of 8 follow-up years.

**Results:** 107,310 (30%) participants had hypertension at UK Biobank enrolment, and 9,969 (3%) developed a pneumonia during follow-up. Prevalent hypertension at baseline was significantly associated with increased risk for incident respiratory disease including pneumonia (hazard ratio 1.36 (95% confidence interval 1.29 to 1.43), P<0.001), acute respiratory distress syndrome or respiratory failure (1.43 (1.29 to 1.59), P<0.001), and chronic lower respiratory disease (1.30 (1.25 to 1.36), P<0.001), independent of age, age^2^, sex, smoking status, BMI, prevalent diabetes mellitus, prevalent coronary artery disease, and principal components of ancestry. Mendelian randomisation analyses indicated that genetic predisposition to a 5 mmHg increase in blood pressure was associated with increased risk of incident pneumonia for SBP (1.08, (1.04 to 1.13), P<0.001) and DBP (1.11 (1.03 to 1.20), P=0.005). Additionally, consistent with epidemiologic associations, increase in blood pressure genetic risk was significantly associated with reduced forced expiratory volume in the first second, forced vital capacity, and the ratio of the two (P<0.001 for all).

**Conclusions:** These results strongly suggest that elevated blood pressure independently increases risk for pneumonia and reduces pulmonary function. Maintaining adequate blood pressure control, in addition to other measures, may reduce risk for pneumonia. Whether the present findings are generalizable to novel coronavirus disease 2019 (COVID-19) require further study.

**Summary Box:** *Section 1: What is already known on this topic:*

- Hypertension has been associated with pneumonia in small observational studies.
- Based on early epidemiologic analyses, hypertension is described as a risk factor for SARS-CoV-2 infection and associated novel coronavirus disease 2019 (COVID-19).
- The influence of hypertension on pneumonia risk is difficult to assess in traditional observational studies.

*Section 2: What this study adds:*

- Our pre-COVID-19 analyses are consistent with a causal relationship between increased blood pressure and increased risk for incident respiratory infections, as well as between increased blood pressure and reduced pulmonary function.
- These results support hypertension as a pneumonia risk factor; efforts to optimize blood pressure may reduce risk for pneumonia.

## Introduction

Hypertension is a highly prevalent, modifiable risk factor for cardiovascular disease and mortality^1^. Epidemiologic analyses have correlated hypertension with pneumonia risk in small studies^2 3^. Whether this represents a direct consequence of hypertension or influence of co-morbid risk factors such as age, diabetes mellitus, air pollution, or smoking is unclear.

Hypertension has emerged as one of the most well-recognized risk factors for developing novel coronavirus disease 2019 (COVID-19) from SARS-CoV-2 infection^4-7^. In particular, patients with COVID-19 who develop acute respiratory distress syndrome (ARDS) are particularly enriched for hypertension^7^. Angiotensin converting enzyme (ACE) is an endogenous regulator of blood pressure that is highly expressed in the lung and has been linked to risk for acute respiratory distress syndrome (ARDS)^8 9^. ACE2, a homolog and endogenous counter-regulator of ACE, is expressed in a subset of type II alveolar cells and is an important facilitator of cell entry by SARS-CoV-2^8-10^. ACE2 has also been implicated in lung injury related to other respiratory viruses and bacterial pathogens^11-13^. While professional societies have encouraged patients to remain on ACE inhibitors during the present pandemic, recommendations were largely made in the absence of robust human data.

Although epidemiologic analyses support a plausible causal relationship between blood pressure regulation and respiratory infection risk, confounding from comorbid conditions limit such inference. Importantly, comorbid factors such as advanced age, diabetes mellitus, and cardiovascular disease have also been described as independent risk factors for pneumonia previously as well as for COVID-19. Mendelian randomisation is a statistical approach using genetic proxies for an exposure as opposed to the exposure itself to mitigate risks for confounding facilitating more robust causal inference^14^. Blood pressure is a highly heritable trait with several known associated genomic loci that may serve as a robust aggregated genetic proxy for Mendelian randomization^15^.

In the UK Biobank prior to the COVID-19 pandemic, we (1) estimate the epidemiologic association of hypertension with incident pneumonia risk and indices of pulmonary function, and (2) apply Mendelian randomisation to test the hypothesis that blood pressure independently causally influences risks for pneumonia and reduced pulmonary function.

## Methods

### UK Biobank

Individual-level genomic data and longitudinal phenotypic data from the UK Biobank, a large-scale population-based cohort with genotype and phenotype data in approximately 500,000 volunteer participants recruited from 2006-2010 was used^16^. Baseline assessments were conducted at 22 assessment centres across the UK using touch screen questionnaire, computer assisted verbal interview, physical tests, and sample collection including for DNA. Additional details regarding the study protocol are described online (www.ukbiobank.ac.uk). Of 488,377 individuals genotyped in the UK Biobank, we used data for 377,143 participants with white British ancestry consenting to genetic analyses, with genotypic-phenotypic sex concordance, without sex aneuploidy, and one from each pair of 1^st^ or 2^nd^ degree relatives selected randomly. Participants provided informed consent as previously described. Secondary use of the data was approved by the Massachusetts General Hospital institutional review board (protocol 2013P001840) and facilitated through UK Biobank Application 7089.

Genome-wide genotyping was previously performed in the UK Biobank using two genotyping arrays sharing 95% of marker content: Applied Biosystems UK BiLEVE Axiom Array (807,411 markers in 49,950 participants) and Applied Biosystems UK Biobank Axiom Array (825,927 markers in 438,427 participants) both by Affymetrix (Santa Clara, CA)^16^. Variants used in the present analysis include those also imputed using the Haplotype Reference Consortium reference panel of up to 39 million single nucleotide polymorphisms (SNPs)^17 18^. Poor quality variants and genotypes were filtered as previously described^16^.

### Phenotypes

#### Hypertension, covariates, and medication measures

Disease definitions for hypertension and clinical disease covariates are as previously described^19^. In brief, hypertension was defined by self-reported hypertension and billing codes for essential hypertension, hypertensive disease with and without heart failure, hypertensive heart and renal diseases, and secondary hypertension. Prevalent coronary artery disease (CAD), was defined by billing codes for heart attack, angina pectoris, unstable angina, myocardial infarction, coronary atherosclerosis, coronary artery revascularization, and other acute, subacute, and chronic forms of ischemic heart disease, or with self-reported angina, heart attack/myocardial infarction, coronary angioplasty +/- stent, or coronary artery bypass graft (CABG) surgery. Diabetes mellitus included billing codes for type 1, type 2, and gestational diabetes mellitus, as well as self-reported diabetes mellitus and insulin use.

Blood pressure medications were characterized by medication type into angiotensin converting enzyme inhibitors (ACEi), angiotensin receptor blockers (ARB), or other medication. The list of ACEi and ARBs considered in UK Biobank participants is provided in **Supplementary Table 1**.

Patients taking at least one ACEi were included in the ACEi category, patients taking no ACEi but at least one ARB were included in the ARB category, and patients who reported that they were taking a blood pressure medication (via UK Biobank Field IDs 6153 and 6177) but were not taking an ACEi or ARB were included in the ‘other’ category.

#### Respiratory outcomes

Clinical disease definitions for our primary outcome (pneumonia) and related respiratory outcomes are detailed in **Supplementary Table 2**. In summary, these included respiratory diseases using the first reported occurrences of respiratory system disorders in Category 2410 as categorized by the UK Biobank (http://biobank.ndph.ox.ac.uk/showcase/label.cgi?id=2410) which maps primary care data, ICD-9 and ICD-10 codes from hospital inpatient data, ICD-10 codes in death register records, and self-reported medical conditions reported at the baseline, to ICD-10 codes. For each set of phenotypes, the time to first incident event after baseline examination in individuals free of prevalent history of respiratory system disorder was used. Pneumonia includes viral, bacterial, and unspecified aetiologies (J12-J18). Influenza or viral pneumonia includes confirmed or suspected influenza or viral pneumonia (J09-J12). The acute upper respiratory infections category includes acute nasopharyngitis, sinusitis, pharyngitis, tonsillitis, laryngitis, tracheitis, croup, epiglottitis, or upper respiratory infections of multiple and unspecified sites (J00-J06). The other acute lower respiratory infections category includes acute bronchitis, bronchiolitis, or unspecified acute lower respiratory infections (J20-J22). The other diseases of the upper respiratory tract category includes rhinitis, nasopharyngitis, pharyngitis, chronic sinusitis, nasal polyps, other disorders of the nose and nasal sinuses, chronic diseases of the tonsils and adenoids, peritonsillar abscess, chronic laryngitis, laryngotracheitis, diseases of the vocal cords and larynx, or other diseases of the upper respiratory tract (J30-J39). Chronic lower respiratory diseases include bronchitis, emphysema, chronic obstructive pulmonary disease, asthma, bronchiectasis (J40-J47). Other interstitial respiratory diseases refer to acute respiratory distress syndrome (ARDS), pulmonary oedema, or other interstitial pulmonary diseases (J80, J81, J84). Respiratory failure refers to J96 (respiratory failure not elsewhere classified).

Quantitative phenotypes included best-measure pulmonary function tests from spirometry using a Vitalograph Pneumotrac 6800 (Buckingham, United Kingdom), including forced expiratory volume in the first second (FEV1), forced vital capacity (FVC), and the ratio of these two measurements (FEV1/FVC). For each individual measurement, extreme outliers were determined and filtered by adjusting the traditional box and whisker upper and lower bounds and accounting for skewness in the phenotypic data identified using the Robustbase package in R (setting range=3) (https://cran.r-project.org/web/packages/robustbase/robustbase.pdf). Phenotypes were then inverse rank normalized to mean 0 and standard deviation (SD) 1 for analysis.

### Statistical methods

#### Association between prevalent hypertension and respiratory conditions in the UK Biobank

Phenotypic association of prevalent hypertension with incident respiratory diseases and with pulmonary function tests was performed using Cox proportional hazards models among individuals without the corresponding prevalent condition in R (version 3.5, R Foundation, Vienna, Austria). The proportional hazards assumption was assessed by Schoenfeld residuals and was satisfied. The sparsely adjusted model included age, age^2^, sex, smoking status (current, previous, or never smoker), and the first ten principal components of ancestry. The fully adjusted model additionally incorporated body-mass index (BMI), prevalent diabetes mellitus, and prevalent coronary artery disease.

#### Genetic instruments for blood pressure and association analyses

One-sample Mendelian randomisation was performed in the UK Biobank by associating inverse-rank normalized systolic blood pressure and diastolic blood pressure polygenic risk scores (SBP PRS, DBP PRS) with incident respiratory disease phenotypes. The 75 variants comprising each genetic instrument (**Supplementary Tables 3, 4**) were determined by identifying genome-wide significant (P<5×10^−8^), largely uncorrelated (linkage disequilibrium r^2^<0.2) variants for each phenotype from the International Consortium for Blood Pressure (ICBP) GWAS summary statistics across 299,024 individuals among 77 cohorts excluding the UK Biobank^15^.

Additive polygenic risk scores were determined as such: 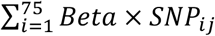, where *Beta* is the weight for each allele of *SNP*_*i*_ from the ICBP GWAS summary statistics, and *SNP*_*Ij*_ is the number of alleles (i.e., 0, 1, or 2) for *SNP*_*i*_ in participant *j* in the UK Biobank. To confirm that the SBP PRS and DBP PRS were strong instruments, we assessed their associations with SBP and DBP, adjusted for blood pressure medications by adding 15 and 10 mmHg to SBP and DBP, respectively, as previously done^20 21^. Each genetic instrument was validated against its exposure by calculating an F-statistic derived from unadjusted linear regression of the exposure against its PRS. An F-statistic greater than 10 indicates low risk of weak-instrument bias.

Additional sensitivity analyses tested associations between each PRS and potential social and lifestyle confounders including the Townsend deprivation index for socioeconomic status estimation^22^, smoking status (Field ID 20116), alcohol intake frequency (Filed ID 1558), vegetable serving intake (Field ID 104060), handfuls of sweet intake (Field ID 102330), significant life stressor over the past two years (Field ID 6145), and exercise frequency (Field ID 3637).

Association analysis of the SBP and DBP PRS with incident respiratory diseases was performed using Cox proportional hazards models in R (version 3.5, R Foundation, Vienna, Austria), adjusting for age, age^2^, sex, smoking status, and the first ten principal components of ancestry. The proportional hazards assumption was assessed by Schoenfeld residuals and was satisfied. Association analyses between the SBP and DBP PRS and pulmonary function tests was performed using a generalized linear model adjusted for the same covariates.

Association of the 75 variants for each PRS with both prevalent and incident pneumonia was performed using a logistic regression Wald test in Hail-0.2, adjusting for age, age^2^, sex, smoking status, the first ten principal components of ancestry, and genotype array. Using these associations, two-sample Mendelian randomization was performed using the ICBP-derived SBP and DBP genetic instruments as exposures and the respective effects of each variant on pneumonia in the UK Biobank as outcomes. Two-sample Mendelian randomization was performed using both the robust, penalized inverse variance weighted (IVW) method from the MendelianRandomization package in R^23 24^, as well as robust adjusted profile score (MR-RAPS)^25^ for comparison. IVW 2-sample Mendelian randomization uses a weighted linear regression of the ratio of the SNP effects on the outcomes to the SNP effects on the risk factor, without using an intercept term. MR-RAPS models the systematic pleiotropy using a random effects model to create a robust adjusted profile score. We additionally use the TwoSampleMR package in R (https://mrcieu.github.io/TwoSampleMR/index.html) to perform multiple sensitivity analyses including heterogeneity tests, the MR Egger intercept test for horizontal pleiotropy, and leave-one-out analyses to determine if there is a single variant driving the genetic association.

## Results

### Baseline characteristics

A total of 377,143 genotyped individuals in the UK Biobank passed quality control criteria. Among these individuals, median age was 58 [interquartile range 51-63] years, 202,369 (53.7%) were female, 18,943 (5.0%) had diabetes mellitus, 107,310 (29.7%) had hypertension, and 20,825 (5.7%) had coronary artery disease. 170,713 (45.4%) of individuals were prior or current smokers, and 85,565 (22.6%) of individuals were on antihypertensive medications (**Supplementary Table 5**).

Across an average follow-up time of 8 years (IQR 7-11 years), 9,969 (2.6%) individuals developed a pneumonia, 11,972 (3.3%) developed influenza or pneumonia, 18,172 (5.5%) developed an acute upper respiratory infection, 21,734 (6.2%) developed other lower respiratory infection (e.g., bronchitis), 12,963 (4.0%) developed a chronic lower respiratory disease, 1,792 (0.5%) developed other interstitial respiratory disease, and 2,621 (0.7%) developed ARDS or respiratory failure.

### Epidemiological association of prevalent hypertension with incident respiratory disease

Prevalent hypertension was independently associated with risk of incident respiratory disease including pneumonia (1.36 (1.29 to 1.43), P<0.001), chronic lower respiratory disease (11.30, (1.25 to 1.36), P<0.001), influenza or pneumonia (1.31 (1.25 to 1.37), P<0.001), ARDS or pulmonary failure (1.43 (1.29 to 1.59), P<0.001), other lower respiratory infections (1.15 (1.37 to 1.45), P<0.001), other acute upper respiratory infection (1.06 (1.11 to 1.19), P=0.002), other interstitial respiratory disease (1.16 (1.02 to 1.31), P=0.022), and influenza or viral pneumonia (1.12 (1.01 to 1.23), P=0.032), after adjustment for age, age^2^, sex, smoking status, BMI, prevalent diabetes, prevalent coronary artery disease, and the first ten principal components of population stratification (**Figure 1**).

**Figure 1:**
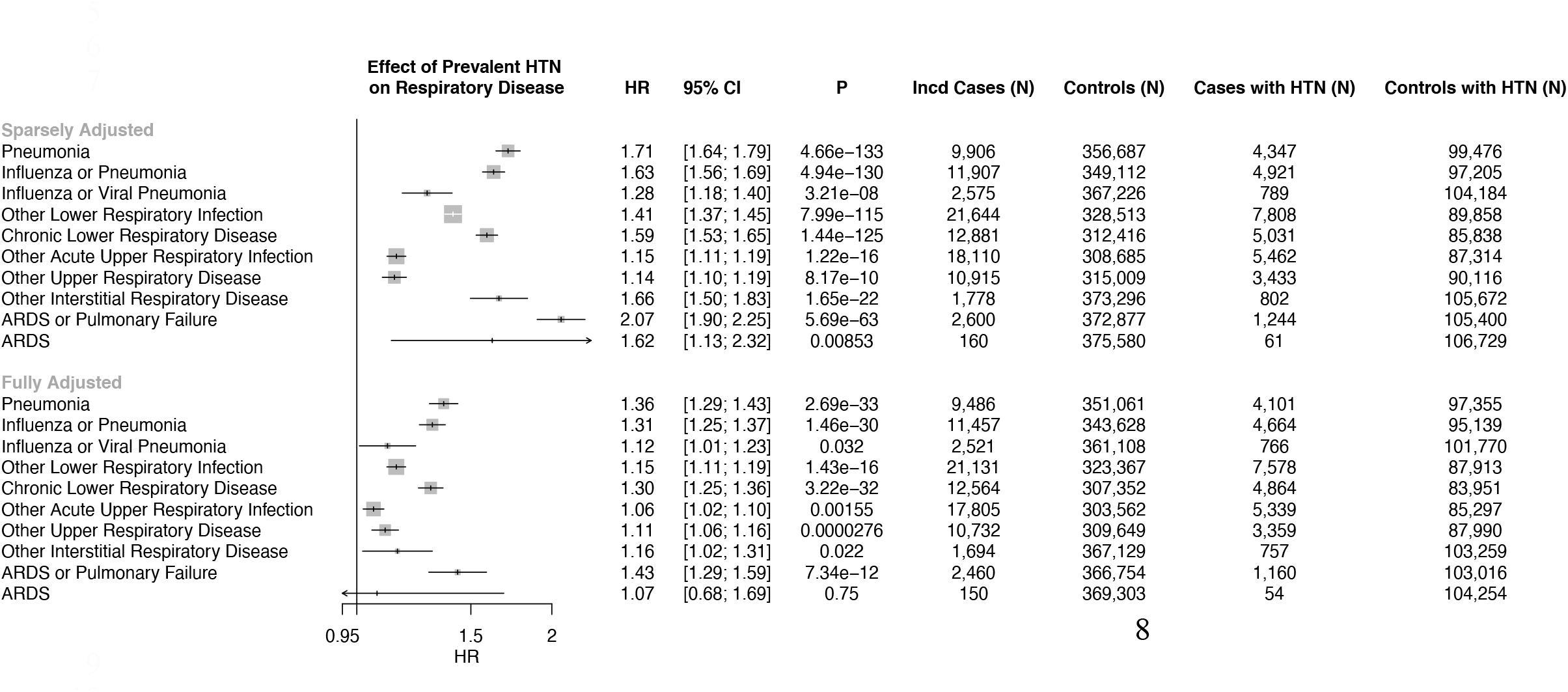
Epidemiological association of prevalent hypertension with respiratory diseases. Association between prevalent hypertension (HTN) and incident respiratory disease is shown in a sparsely adjusted and fully adjusted model. The sparsely adjusted model is adjusted by age, age^2^, sex, smoking status (current, prior, or never smoker), and the first ten principal components of population stratification. The fully adjusted model is additionally adjusted by prevalent coronary artery disease, prevalent diabetes, and body mass index. HTN = Hypertension, ARDS = Adult respiratory disease syndrome, HR = hazard ratio

### Epidemiological association of antihypertensive use with incident pneumonia

Among the participants, ACEi were prescribed for 37,865 (10.0%), ARBs for 13,207 (3.5%), and other antihypertensives for 34,493 (9.1%). Prescriptions for ACEi, ARBs, or other antihypertensive medicines were similarly associated with increased risk for incident pneumonia (**Figure 2**). However, upon further adjustment for prevalent hypertension status, we did not observe an association of these medication classes with risk for pneumonia.

**Figure 2:**
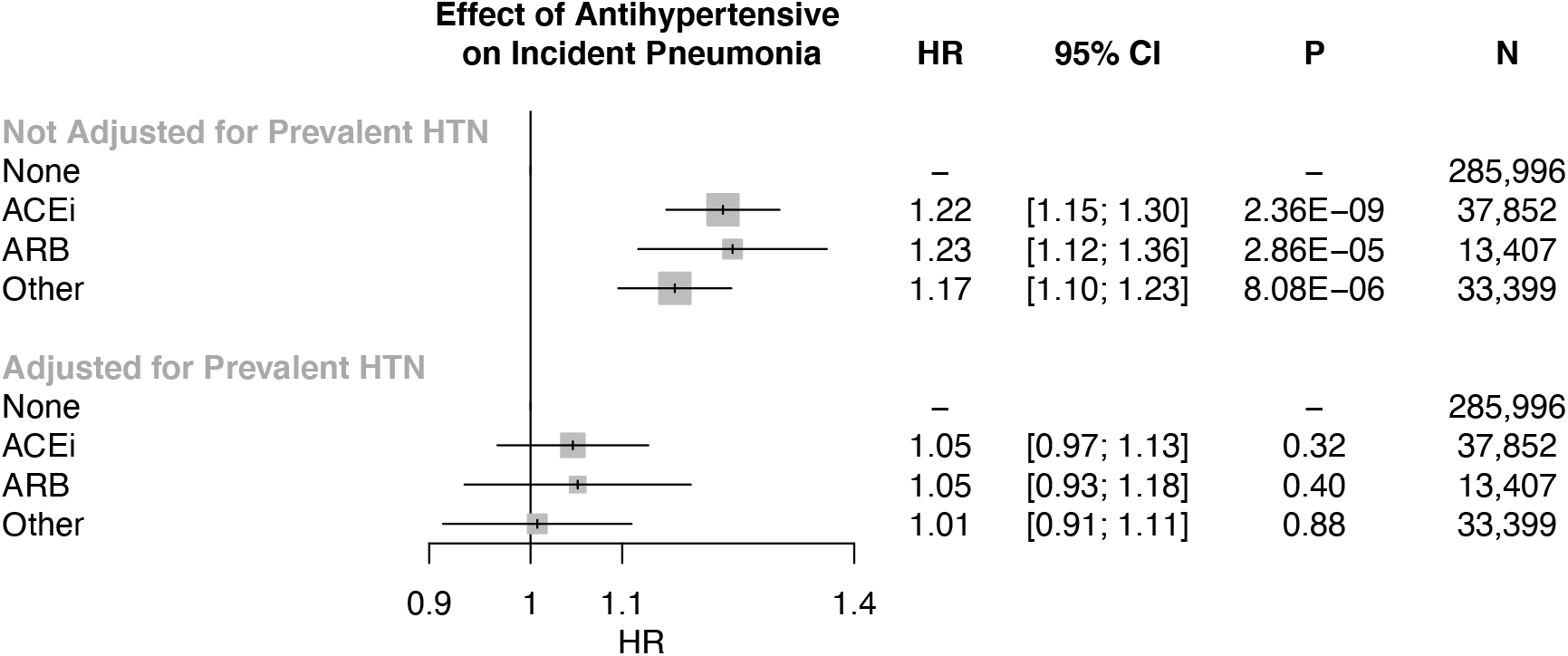
Epidemiological association of antihypertensive use with incident pneumonia. Association of antihypertensive use with incident pneumonia, adjusted by age, age2, sex, smoking status, prevalent coronary artery disease, prevalent diabetes, body mass index, and the first ten principal components of population stratification, displayed with and without adjusting for prevalent hypertension, suggests that the effect of antihypertensives on increased risk of incident pneumonia is driven by hypertensive status. ACEi = angiotensin converting enzyme inhibitor, ARB = angiotensin receptor blocker, HR = hazard ratio

### Genetic association of blood pressure with incident respiratory disease

We first used one-sample Mendelian randomisation to determine whether a genetic predisposition to increased blood pressure is associated with increased risk for incident pneumonia as well as other respiratory diseases.

Our SBP and DBP genetic instruments each consisted of 75 independent variants (linkage disequilibrium r^2^<0.2) that were genome-wide significant among 299,024 individuals external to the UK Biobank from the International Consortium for Blood Pressure Genomics. The resulting PRS for SBP and, separately, for DBP were significantly associated with their respective phenotypes in the UK Biobank, with each SD increase in the SBP PRS increasing SBP by 2.26mmHg (F-statistic: 4176) and each SD increase in the DBP PRS increasing DBP by 1.32 mmHg (F-statistic: 4785) (**Supplementary Table 6**). 53 SNPs (or SNPs in perfect linkage disequilibrium) were common to both SBP and DBP PRS. Sensitivity analyses were performed to assess for potential social and lifestyle confounders associating with the SBP and DBP PRS, and found no significant associations between the PRS and the Townsend deprivation index for socioeconomic status estimation, smoking status, alcohol intake frequency, vegetable intake, sweet intake, significant life stressor in the past two years, or exercise frequency (**Supplemental Table 7**).

Each SD increase in the SBP PRS was associated with a significant increased risk of incident pneumonia (1.04 (1.02 to 1.06), P<0.001), influenza or pneumonia (1.03 (1.01 to 1.05), P=0.003), and other lower respiratory infection (1.02 (1.00 to 1.03), P=0.029). Each SD increase in the DBP PRS was also associated with an increased risk of incident pneumonia (1.03, (1.01 to 1.05), P=0.005), and other lower respiratory infections (1.02 (1.00 to 1.03), P=0.012) (**Figure 3**). Accordingly, each 5mmHg increase in SBP conferred by the SBP PRS was associated with a significant increased risk of incident pneumonia incident pneumonia (1.08 (1.04 to 1.13), P<0.001), influenza or pneumonia (1.06 (1.02 to 1.11), P=0.003), and other lower respiratory infection (1.03 (1.00 to 1.06), P=0.029). Additionally, each 5mmHg increase in DBP conferred by the DBP PRS was associated with an increased risk of incident pneumonia (1.11, (1.03 to 1.20), P=0.005), and other lower respiratory infections (1.07 (1.01 to 1.12), P=0.012).

**Figure 3:**
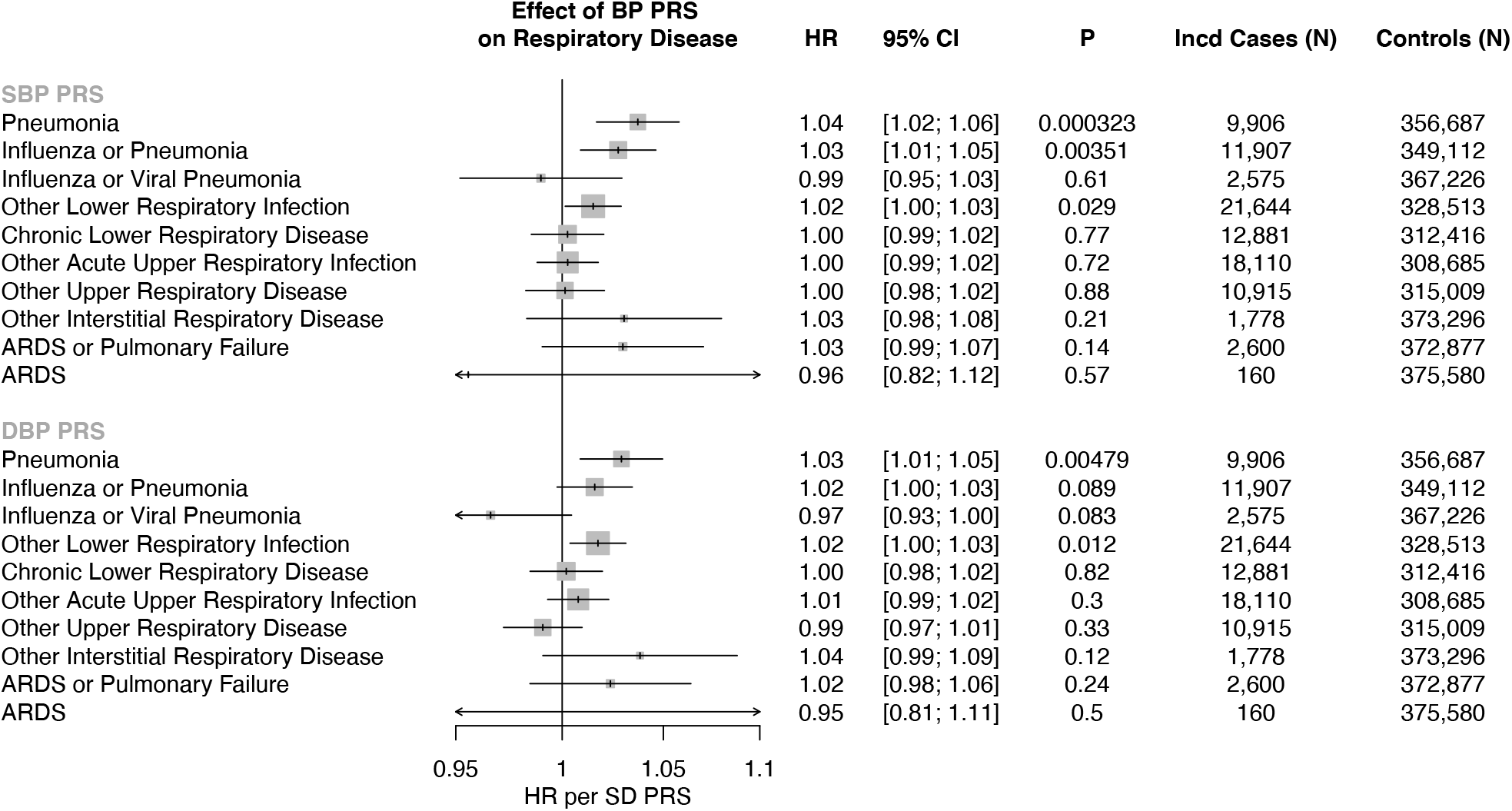
Genetic association of blood pressure with incident respiratory disease. Association of systolic blood pressure polygenic risk score (SBP PRS) and diastolic blood pressure polygenic risk score (DBP PRS) with incident respiratory disease, adjusted for age, age^2^, sex, smoking status, and the first ten principal components of population stratification in the UK Biobank. Effects are interpreted as hazard ratio (HR) per standard deviation (SD) increase of the respective PRS. Of note, each SD increase in the SBP PRS increases SBP by 2.26mmHg, and each SD increase in the DBP PRS increases DBP by 1.32 mmHg in the UK Biobank. SBP PRS = systolic blood pressure polygenic risk score, DBP PRS = diastolic blood pressure polygenic risk score, HR = hazard ratio, SD = standard deviation

Two-sample Mendelian randomization was additionally performed using pneumonia association statistics, from the UK Biobank, for the 75 variants, with blood pressure summary statistics external to the UK Biobank as above. Both penalized, robust, inverse-variance weighted (IVW) and robust adjusted profile score (MR-RAPS)^25^ methods produced similarly significant results (**Supplementary Table 8**). Sensitivity analyses were additionally performed to analyse the robustness of the results. Across both SBP and DBP genetic instruments, the IVW heterogeneity test and the MR Egger regression intercept term are both insignificant, suggesting negligible contribution of heterogeneity and directional horizontal pleiotropy (**Supplementary Table 8**). Additionally, the Steiger directionality test^26^ suggests the correct causal direction of blood pressure on pneumonia. Lastly, leave-one-out analyses was additionally performed, suggesting that no single SNP drives the observed association (**Supplementary Figure 1)**. Variants previously described to influence *ACE* expression in the lung (rs145126552, rs4277405) or kidney (rs6504163) in GTEx^27^, were not individually associated with pneumonia risk (P>0.05).

### Epidemiological association of prevalent hypertension with pulmonary function tests

Secondary analyses identified significant associations between prevalent hypertension and reduced pulmonary function tests (FEV1: -0.088 SD (-0.09 to -0.08), P<0.001; FVC: -0.85 SD (- 0.09 to -0.08), P<0.001; FEV1/FVC: -0.013 SD (-0.02 to -0.01), P<0.001), independent of age, age^2^, sex, smoking status, body mass index, prevalent coronary artery disease, prevalent diabetes mellitus, and the first ten principal components of ancestry (**Figure 4A**).

**Figure 4:**
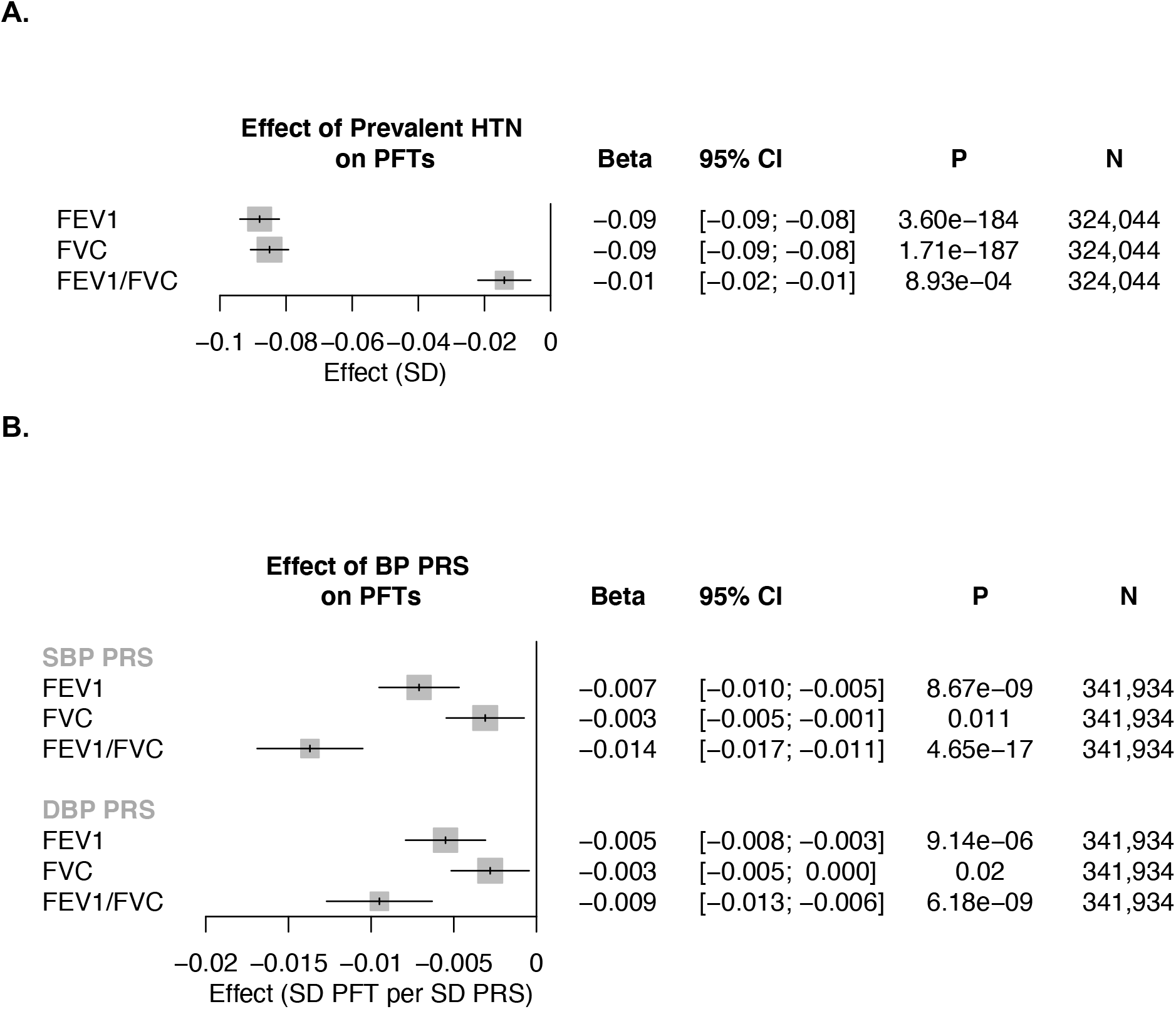
Epidemiological and genetic association of elevated blood pressure with pulmonary function tests. **A)** Association of prevalent hypertension with pulmonary function tests (PFT), adjusted for age, age2, sex, smoking status, prevalent coronary artery disease, prevalent diabetes, body mass index, and the first ten principal components of population stratification in the UK Biobank. **B)** Association of systolic blood pressure polygenic risk score (SBP PRS) and diastolic blood pressure polygenic risk score (DBP PRS) with pulmonary function tests (PFT), adjusted for age, age2, sex, smoking status, and the first ten principal components of population stratification in the UK Biobank. Effects are interpreted as SD change in PFT per SD increase of the respective PRS. Of note, each SD increase in the SBP PRS increases SBP by 2.26mmHg, and each SD increase in the DBP PRS increases DBP by 1.32 mmHg in the UK Biobank. FEV1 = Forced expiratory volume in 1-second, FVC = Forced vital capacity, HTN = hypertension, PFT = pulmonary function test

### Genetic association of blood pressure with pulmonary function tests

Each SD increase in SBP PRS was significantly associated with reduced pulmonary function tests (FEV1: -0.007 SD (-0.01 to -0.005), P<0.001; FVC: -0.003 SD (-0.005 to -0.001), P=5.91×10^−3^; FEV1/FVC: -0.014 SD, (-0.017 to -0.011), P<0.001). Similar associations were identified for between DBP PRS and pulmonary function tests (FEV1: -0.005 SD(-0.008 to - 0.003), P<0.001; FVC: -0.003 SD (-0.005 to 0), P=0.02; FEV1/FVC: -0.009 SD, (-0.013 to - 0.006), P<0.001) (**Figure 4B**).

## Discussion

In a large, prospective, population-based cohort, we show that prevalent hypertension is a risk factor for incident pneumonia, lower respiratory infections, ARDS or respiratory failure, as well as many other respiratory diseases. Additionally, our epidemiological analyses also demonstrate an association between prevalent hypertension and increased pulmonary obstruction as indicated by reduced FEV1/FVC. Our Mendelian randomisation studies imply that the relationship between increased blood pressure with increased risk for pneumonia as well as reduced pulmonary function may be causal.

Our findings may have important implications for the prevention of pneumonia. First, our study establishes that hypertension is an important independent risk factor for the development of pneumonia. Our observation is consistent with prior unadjusted estimates in retrospective analyses^2 28^. Although significantly associated, the effect of hypertension adjusted for important confounders on future risk for pneumonia is more moderate, as anticipated. Furthermore, our genetic analyses are consistent with a causal relationship between blood pressure and pneumonia. Hypertension has been proposed to promote key factors which may predispose to infection via several potential mechanisms. (1) Hypertensive stimuli promote dysregulation of the adaptive immune response. Chronic angiotensin II infusions in mice increase markers of T lymphocyte activation and perivascular adipose infiltration^29^. Additional murine studies have indicated that monocytes and neutrophils may be key factors in angiotensin II-mediated hypertension and resultant vascular dysfunction^30 31^. A recent Mendelian randomisation study supported a causal relationship between blood pressure and subsequent alteration in neutrophil, monocyte, and eosinophil indices^32^. (2) Endothelial dysfunction as a consequence of hypertension may promote infection. Dysregulation of nitric oxide release and signalling in murine models of pulmonary inflammation leads to exacerbated lung injury^33^. Furthermore, superoxides generated from endothelial NADPH oxidase may play a role in exacerbating influenza-associated lung injury in experimental models^34^.

Second, in addition immunologic and vascular effects, blood pressure elevations may result in pulmonary function alterations predisposing to the development of pneumonia. Our Mendelian randomisation analyses with pulmonary function tests support a causal association between increased blood pressure and indices of increased pulmonary obstruction as indicated by reduced FEV1/FVC. Several prior studies have linked hypertension with decreased performance on pulmonary function tests, reduced lung function, and increased pulmonary obstruction^35-37^. Chronic obstructive pulmonary disease (COPD) is a well-established risk factor for pneumonia, and is co-morbid with several cardiovascular diseases and risk factors including hypertension^38^. Our study extends these observations to show that increased blood pressure may causally lead to increased pulmonary obstruction representing a putative mechanism toward heightened pneumonia risk. Together, these studies and others suggest several mechanisms which may link hypertension and pulmonary obstruction: 1) both may involve physiological degradation of arterial and airway elasticity^37^, 2) endothelial and vascular dysfunction may also influence pulmonary vascular endothelial cells and lead to pulmonary vascular dysfunction resulting in lung tissue destruction and airway obstruction^40^, and 3) systemic inflammation associated with hypertension may additionally alter pulmonary function^41 32^.

Third, since our results are consistent with a causal relationship between blood pressure and pneumonia risk, blood pressure optimization is anticipated to reduce pneumonia risk in the population orthogonal to other strategies aimed at reducing infection risk. Since *ACE* is highly expressed in the lung with adverse effects, such as cough, there were initial concerns about ACEi potentially exacerbating pulmonary diseases or leading to pulmonary infections. The observational association of antihypertensives with increased pneumonia risk is rendered nonsignificant when adjusting for hypertension consistent with confounding by indication. A meta-analysis comprising of randomized controlled and observational studies indicated that ACEi may be protective for pneumonia risk^42^. As detailed earlier, since ACE2 may serve as a receptor for respiratory viral entry and is an endogenous counter-regulator of ACE, conflicting hypotheses for ACEi and COVID-19 risk have emerged^43-45^. We do not observe that genetic variants influencing the expression of *ACE* in the lung or kidney significantly increase the risk of pneumonia. Whether genetic variants at *ACE* have outsized influence on COVID-19 risk requires further study. However, our results are aligned with current recommendations to maintain stable, normal blood pressure, including with ACEi as indicated^46 47^. Consistent with this finding, the Systolic Blood Pressure Intervention Trial (SPRINT) also showed that intensive blood pressure reduction (to a mean SBP of 121 mmHg) resulted in fewer cases of incident pneumonia (2.1% versus 2.4%) compared to standard blood pressure reduction (to a mean SBP of 136 mmHg)^48^. Additionally, a separate multi-center study of 1,128 patients with hypertension diagnosed with COVID-19 showed that inpatient use of ACEi/ARB was associated with lower risk of all-cause mortality compared with ACEi/ARB non-users (HR 0.37, P=0.03)^49^. Thus, withdrawal of ACEi and challenges of blood pressure management while promoting physical distancing, may not only have untoward cardiovascular consequences but may inadvertently increase pneumonia risk.

### Strengths and limitations

This study has several strengths, including the analysis of a large, genotyped population-based cohort with high fidelity phenotyping, including with subclinical respiratory phenotypes. Furthermore, diverse phenotyping facilitates extensive individual-level covariate adjustment in the models and sensitivity analyses to assess for pleiotropy. Our overall study design, with the incorporation of Mendelian randomisation, permits more robust causal inference in humans beyond observational prospective analyses. While our study has several strengths, some limitations should be considered. First, the present analyses were conducted among individuals of white British ancestry residing in the UK; whether the present findings generalize to diverse ethnicities and other geographic regions remains to be tested. Second, our model assumes the association of the genetic instrument to the outcome occurs via the primary exposure and is not confounded by pleiotropy^50 51^. We have performed a number of sensitivity analyses to assess for possible confounders, in particular, we observe that our genetic instruments for blood pressure are not associated with key socioeconomic and lifestyle factors influencing both blood pressure and pneumonia risk. We further maintain a sparsely adjusted model in our one-sample Mendelian randomization analyses to reduce the potential for collider bias^52^. Additionally, secondary 2-sample Mendelian randomization analyses provide consistent results with no evidence of horizontal pleiotropy, heterogeneity, or individual variants driving the association. Third, while our results do not yield an association between *ACE* eQTLs and pneumonia risk, we cannot rule out the possibility of reduced power for this analysis. Fourth, outcomes for the present analyses occurred prior to the COVID-19 pandemic. The first release of COVID-19 phenotypes from the UK Biobank participants has few COVID-19 events (approximately 0.1%), which limits power. Furthermore, with testing data only in 0.3% of UK Biobank participants, analyses are currently limited by ascertainment bias. Whether the current findings translate to COVID-19 require further verification.

## Conclusions

Our study provides evidence that hypertension directly influences the risk of pneumonia. Vigilance for blood pressure monitoring and management may reduce risk for pneumonia, including potentially for the ongoing COVID-19 pandemic.

## Data Availability

All of the individual-level data used in the manuscript are available by application through the UK Biobank (https://www.ukbiobank.ac.uk).

## Acknowledgments

The authors would like to acknowledge and thank the participants and staff of the UK Biobank and of the ICBP consortium cohorts. This work was supported by UK Biobank application number 7089.

